# Cardiovascular hospital demand projections for Suriname up to 2050 with implications for health system planning

**DOI:** 10.64898/2026.11.08.26367566

**Authors:** Ritesh A. Dhanpat, Rakesh Gajadhar Sukul, Jerry R. Toelsie, Marina Bersaoui, Angele Mendeszoon, Vanita Jairam, Robbert Bipat

## Abstract

This study developed a data-driven decision-support framework to project annual hospital admissions and in-hospital mortality for ischemic heart disease (IHD) and cerebrovascular disease in Suriname from 2023 through 2050.

Using retrospective discharge data from 2013–2022 across three major public hospitals—accounting for approximately 71.8% of national admissions—historical trends were analyzed using log-linear regression across six age groups. Population projections relied on the UN World Population Prospects 2024 Revision. Future admissions and deaths were modelled across five policy pathways: status quo, current HEARTS, strengthened HEARTS, full HEARTS targets, and full HEARTS combined with improved hospital survival, with uncertainty assessed via 10,000 Monte Carlo simulations.

Results showed that admission rates increased for most age groups between 2013 and 2022. Under the status quo pathway, combined cardiovascular admissions are projected to reach 15,592 in 2030, 30,772 in 2040, and 62,643 in 2050. Compared to the status quo in 2050, projected admissions would drop to 58,073 under current HEARTS, 41,690 under strengthened HEARTS, and 27,469 under full HEARTS—representing median reductions of 6.0%, 33.5%, and 56.1%, respectively. Additionally, projected 2050 deaths would decrease from 2,644 under the status quo to 1,155 under full HEARTS, and further to 924 with improved hospital survival.

Cardiovascular hospital demand could rise substantially if historical trends continue, impacting health-system capacity. While not causal estimates or precise forecasts, these modelled pathways illustrate how routine health data can create a transparent framework to guide cardiovascular prevention, resource allocation, and health-system planning in Suriname.

## Introduction

Cardiovascular disease remains one of the major public health challenges facing countries throughout the Region of the Americas(1). Its consequences extend beyond premature mortality and morbidity to increasing demands on health services, including emergency care, inpatient services, diagnostic capacity, medicines, specialist care and rehabilitation(2). These pressures are particularly important in health systems where financial; workforce and infrastructure constraints limit the ability to expand services in response to rapidly increasing demand(2).

Ischemic heart disease (IHD) and cerebrovascular disease are major contributors to cardiovascular morbidity and mortality. Both are strongly associated with modifiable risk factors, particularly hypertension, diabetes, tobacco use, obesity and physical inactivity. Hypertension is especially important because it contributes to both IHD and cerebrovascular disease and can be addressed through population-level prevention and strengthened primary care(3).

For health-system decision-makers, however, documenting current disease burden is only one part of the policy problem. Historical surveillance describes what has happened, but does not directly indicate how hospital demand may evolve under different future policy trajectories. A health system therefore needs evidence not only about the magnitude and direction of current disease trends, but also about the potential consequences of alternative choices regarding prevention, primary care and hospital services(4).

Suriname provides an important setting in which to examine this issue. Previous analyses of hospital discharge data from the country’s major public hospitals demonstrated sustained increases in cardiovascular and renal disease burden between 2013 and 2022(5). The observed increase in cardiovascular hospital utilization occurred in parallel with persistent exposure to cardiovascular risk factors and continuing demands on a health system with limited specialist and hospital capacity(6).

At the same time, Suriname has begun strengthening cardiovascular prevention through the HEARTS initiative(2). HEARTS in the Americas promotes standardized cardiovascular risk management, evidence-based hypertension treatment, access to essential medicines and technologies, team-based care, and systematic monitoring(7–9). The Surinamese implementation program has expanded substantially, but early implementation data have also identified challenges related to continuity of care, workforce capacity, infrastructure, medicines and financing. The implementation study documented 56 participating facilities, 460 healthcare workers and 11,187 patients, while 52.6% of patients had at least one follow-up visit and blood-pressure control among patients with follow-up measurements reached 56.2% (10). These developments create a policy-relevant question: *how might future cardiovascular hospital demand differ if current trends continue compared with alternative trajectories involving progressively stronger cardiovascular prevention and improved hospital care?*

Scenario modelling can provide one approach to answering this question. By combining routinely collected hospital data, population projections and program implementation parameters, epidemiological evidence can be translated into alternative future trajectories that can be compared at relevant planning horizons.

The objective of this study was therefore to develop a data-driven decision-support framework to project annual IHD and cerebrovascular disease hospital admissions and in-hospital mortality in Suriname from 2023 through 2050. We additionally examined alternative policy pathways representing progressively stronger cardiovascular prevention through HEARTS and improved hospital survival. The projections for 2030, 2040 and 2050 were selected as decision horizons to illustrate the potential implications of alternative policy trajectories for long-term health-system planning and resource allocation.

## Methods

### Study design and setting

We conducted a retrospective hospital-based time-series study followed by model-based projections of IHD and cerebrovascular disease hospital admissions and in-hospital mortality. The historical analysis used annual hospital discharge data from three major public hospitals in Suriname covering 1 January 2013 through 31 December 2022. These hospitals collectively accounted for approximately 71.8% of all hospital admissions in Suriname during the study period. The 71.8% figure represents overall hospital activity and should not be interpreted as disease-specific national coverage. Acute IHD is predominantly managed at the major hospitals receiving patients with acute cardiovascular presentations. Cerebrovascular disease admissions are concentrated at the largest referral hospital, although additional patients are admitted to other private hospitals. The participating hospitals therefore provide a substantial and clinically relevant source of hospital data for modelling cardiovascular hospital demand, but the dataset does not constitute a complete national registry of all IHD or cerebrovascular disease events.

### Hospital admissions and disease definitions

Hospital admissions were identified from discharge diagnoses recorded in the hospital databases. IHD was defined using ICD-10 codes I20–I25. Cerebrovascular disease was defined using ICD-10 codes I60–I69, consistent with the disease definitions used in the preceding analyses of the research program. Because the I60–I69 category includes I69, which represents sequelae of cerebrovascular disease, the operational outcome is referred to as cerebrovascular disease rather than stroke throughout this manuscript. The analysis used six age groups: 0–29, 30–39, 40–49, 50–59, 60–69, and ≥70 years.

The unit of analysis was the hospital admission, rather than the individual patient. Patients with multiple admissions during the study period could therefore contribute more than one observation. Unique patients were not identified or deduplicated. Transfers between participating hospitals could also have been recorded as separate admissions and may therefore have resulted in double counting of a single clinical episode.

Discharge status was available for all but 28 admissions. These 28 admissions with unknown discharge diagnosis were excluded from further analysis. Accordingly, the analysis represents recorded hospital admissions with an available discharge status rather than unique patients or incident disease events.

### Data-driven decision-support framework

The projection model was developed as a decision-support framework rather than as a conventional predictive forecasting model. The framework linked three components [1] observed historical hospital admission trends; [2] projected population change; and [3] alternative assumptions regarding cardiovascular prevention and hospital survival. The purpose was to translate routinely collected epidemiological data into alternative future trajectories that could be compared at clinically and operationally relevant planning horizons.

The primary outputs were annual hospital admissions and in-hospital deaths. Results were summarized for 2030, 2040 and 2050 because these years represent near-, medium- and long-term planning horizons. The framework was intended to support policy deliberation rather than determine a single optimal policy. It does not incorporate intervention costs, budget constraints, cost-effectiveness, workforce feasibility or competing health priorities.

### Historical admission-rate estimation

For each age group (a) and disease (d), annual admission rates from 2013–2022 were modelled using log-linear regression: ln *R*_(*a*,*d*,*t*)_ = *β*_0(*a*,*d*)_ + *β*_1(*a*,*d*)_(*t*) where R_a,d,t_ denotes the annual admission rate for age group a, disease d, and year t. The estimated coefficient for calendar year was converted to an annual percentage change (APC) using the formula APC = (*e*^β_1_^ ― 1) × 100 with corresponding 95% CIs derived from the standard error of the estimated regression coefficient. All annual observations available within the historical period were used for estimation. The resulting APC estimates represented the annual multiplicative change in admission rates under the fitted log-linear model.

### Population projections

Population estimates for 2023–2050 were derived using annual population growth rates projected by the United Nations Population Division. The annual rates were obtained from the United Nations World Population Prospects 2024 Revision(11). The projected annual population growth rate declined from 0.909% in 2023 to 0.264% in 2050. The annual rates were applied sequentially to the population estimates to derive annual population values through 2050. Because the UN annual population growth rates used in the analysis were not age-specific, the same annual growth rate was applied to each of the six age groups in each projection year. The resulting age-specific population values therefore represent model-derived estimates based on UN-projected annual population growth rather than directly extracted age-specific UN population projections.

### Projection of hospital admissions

For each age group and disease, annual hospital admissions were projected sequentially from the 2022 baseline. The final implementation of the model applied the estimated annual disease-specific admission trend together with the annual population growth rate: *A*_(*a*,*d*,*t*)_ = *A*_(*a*,*d*,*t*―1)_ ∗ (1 + *APC*_(*a*,*d*)_(1 + *g_t_*_)_, where A_(a,d,t)_ is the number of admissions in age group a, disease d, and year t; APC_(a,d)_ is the estimated annual percentage change; and g_t_ is the UN-projected annual population growth rate.

The age- and disease-specific estimates were summed to obtain total annual hospital demand. All projected values represent conditional trajectories under specified model assumptions, rather than predictions of the exact future number of admissions.

### Projection of in-hospital mortality

For the primary status quo projection, age- and disease-specific case-fatality rates (CFRs) observed in 2022 were held constant throughout the projection period. Projected deaths were calculated as: *Deatℎs_(a,d,t)_ = Admissions_(a,d,t)_ × CFR*_(*a,d*,2022)_

This provided a transparent baseline in which future changes in hospital survival were not assumed unless explicitly incorporated into a scenario. Because the COVID-19 pandemic may have affected hospital utilization, admission patterns, and outcomes during 2020–2022, its potential influence on the long-term trend estimates was assessed separately through a COVID-exclusion sensitivity analysis.

### Policy pathways

Five policy-relevant pathways were modelled.

### Pathway 1: Status quo

Historical admission trends and projected population growth continued without an additional HEARTS-related effect. Age- and disease-specific CFRs remained at 2022 levels.

### Pathway 2: Current HEARTS implementation

This pathway represented the current level of HEARTS implementation. It assumed [1] 80% program reach; [2] 52.6% treatment/follow-up retention; and [3] 56.2% blood-pressure control among patients retained in follow-up. These assumptions resulted in modelled reductions in admissions of 7.5% for IHD and 10.8% for cerebrovascular disease. The 80% program reach was a modelling assumption, not an observed estimate of national HEARTS coverage. The 52.6% follow-up and 56.2% control estimates were derived from available implementation data (10). Because reporting was incomplete across participating facilities, they were not interpreted as nationally representative population estimates.

### Pathway 3: Strengthened HEARTS implementation

This pathway retained 80% program reach and 56.2% blood-pressure control while increasing treatment/follow-up retention to 80%. The resulting modelled reduction in admissions was 35.6% for both IHD and cerebrovascular disease.

### Pathway 4: Achievement of predefined HEARTS targets

This pathway assumed [1] 80% program reach; [2] 80% treatment/follow-up retention; and [3] achievement of the predefined HEARTS target for blood-pressure control. The resulting modelled reduction in admissions was 58.9% for both IHD and cerebrovascular disease.

### Pathway 5: Full HEARTS plus improved hospital survival

This pathway retained the admission reductions of Pathway 4 and additionally applied a 20% reduction in age- and disease-specific CFRs. The 20% CFR reduction was treated as an illustrative modelling assumption, not an empirically established effect of HEARTS or improved hospital care in Suriname. The pathways therefore represent counterfactual policy trajectories, not causal estimates of intervention effectiveness.

### External evidence used to parameterize prevention effects

The relationship between population hypertension control and cardiovascular mortality was parameterized using the ecological analysis by Martinez and colleagues, which examined 36 countries in the Americas(12). The study reported associations between population hypertension control and mortality from IHD and stroke. The reported associations were not specific to Suriname, HEARTS, hospital admissions, or in-hospital mortality. Accordingly, the external estimates were used as model parameters within a counterfactual scenario framework rather than interpreted as direct causal estimates of the effect of HEARTS on hospital admissions.

The transfer from population-level hypertension control to hospital admissions represents an important structural assumption of the model. The scenarios were modelled using a baseline population hypertension-control rate of 21%(13). Because the source relationship was log-linear, the effect was compounded multiplicatively: *Reduction* = 1 ― (1 ― *r*)^Δ*percentage*―*point*^

### Probabilistic sensitivity analysis

Parameter uncertainty was assessed using 10,000 Monte Carlo simulations. Uncertainty was propagated through the stochastic parameters of the final model, including the estimated age- and disease-specific temporal trends and mortality parameters according to the final model specification. The same parameter draws were applied across all five policy pathways within each simulation. This paired approach ensured that comparisons between pathways were based on the same underlying sampled epidemiological parameters. For each outcome and projection year, the median and 2.5th and 97.5th percentiles were calculated and reported as 95% uncertainty intervals (UIs). A fixed random seed of 42 was used to facilitate reproducibility. The population growth trajectory was fixed to the UN-projected annual rates in the final probabilistic analysis and therefore does not contribute stochastic uncertainty to the reported 95% UIs.

### One-way sensitivity analysis

A deterministic one-way sensitivity analysis (OWSA) was conducted by varying individual model parameters across prespecified plausible ranges while holding all other parameters constant. The purpose was to identify parameters that exerted the greatest influence on projected admissions and deaths. The OWSA was interpreted as a deterministic parameter sensitivity analysis and was distinguished from the probabilistic uncertainty analysis.

### Temporal holdout assessment

An internal temporal holdout assessment was performed by fitting the trend models using 2013–2019 data and estimating outcomes for 2020–2022. This assessment was intended to evaluate temporal correspondence between observed and model-estimated values. It was considered an internal temporal assessment, rather than external validation.

### COVID-19 exclusion analysis

Because the COVID-19 pandemic may have disrupted hospital utilization patterns, a sensitivity analysis was performed in which the historical trend models were estimated using 2013–2019 data only. The resulting projections were compared with those from the primary model to assess the influence of the 2020–2022 period on long-term projections.

### Statistical reporting and milestone years

The primary planning outputs were annual admissions and in-hospital deaths in 2030, 2040 and 2050. Scenario differences were calculated within each Monte Carlo simulation before summary statistics were generated. Consequently, uncertainty intervals for scenario reductions represent paired simulation-based comparisons rather than arithmetic subtraction of independently summarized medians. All analyses were conducted using the final reproducible model implementation.

### Ethics

Ethical approval for the hospital-data analyses was obtained from the Commissie Mensgebonden Onderzoek (CMWO), Suriname, letter CMWO-14, dated 25 March 2026. The study used anonymized routinely collected health data, and informed consent was waived in accordance with the applicable national framework.

## Results

### Historical cardiovascular hospital trends

Admission rates for both IHD and cerebrovascular disease increased across most age groups during 2013–2022. For IHD, APCs ranged from 2.56% (95% CI −2.61 to 8.02) among people aged 0–29 years to 9.25% (95% CI 5.36–13.28) among those aged 30–39 years. For cerebrovascular disease, APCs ranged from 2.38% (95% CI −1.96 to 6.91) among people aged 40–49 years to 12.73% (95% CI 3.65–22.61) among those aged 30–39 years. The confidence interval included zero for IHD among people aged 0–29 years and for cerebrovascular disease among people aged 40–49 years. The remaining APC estimates were positive with confidence intervals excluding zero. The magnitude of the observed increases among younger adults indicates that the future cardiovascular hospital burden is not attributable solely to population ageing.

**Table 1.** Historical annual percentage changes in hospital admission rates, Suriname, 2013–2022.

| Age group | IHD APC, % (95% CI) | Cerebrovascular disease APC, % (95% CI) |
|---|---|---|
| 0–29 | 2.56 (−2.61 to 8.02) | 7.01 (1.77 to 12.53) |
| 30–39 | 9.25 (5.36 to 13.28) | 12.73 (3.65 to 22.61) |
| 40–49 | 6.75 (4.51 to 9.03) | 2.38 (−1.96 to 6.91) |
| 50–59 | 5.75 (3.78 to 7.75) | 4.24 (0.91 to 7.68) |
| 60–69 | 5.61 (4.30 to 6.94) | 5.69 (0.69 to 10.95) |
| ≥70 | 6.22 (2.95 to 9.59) | 6.16 (3.18 to 9.24) |

### Cardiovascular hospital demand under the status quo

The status quo pathway demonstrated substantial growth in annual cardiovascular hospital demand over the projection period. Combined annual IHD and cerebrovascular disease admissions were projected at 15,592 in 2030; 30,772 in 2040; and 62,643 in 2050. The probabilistic 2050 status quo estimate was 62,643 admissions (95% UI 46,351–102,113). The increasing uncertainty at longer projection horizons reflects the compounding effect of uncertainty in the underlying age- and disease-specific temporal trends. The projections therefore indicate that continued historical trends could create a substantial increase in cardiovascular hospital demand, even without assuming further deterioration in population health.

### Alternative policy pathways and hospital demand

The divergence between policy pathways increased over time. In 2030, projected total annual admissions were 15,592 under status quo; 14,500 under current HEARTS; 10,352 under strengthened HEARTS; and 6,836 under full HEARTS. In 2040, projected admissions were 30,772 under status quo; 28,516 under current HEARTS; 20,367 under strengthened HEARTS; and 13,408 under full HEARTS. By 2050, projected admissions were 62,643 under status quo; 58,073 under current HEARTS; 41,690 under strengthened HEARTS; and 27,469 under full HEARTS. The full HEARTS and full HEARTS plus improved hospital survival pathways had identical admission trajectories because the additional hospital-care component in Pathway 5 affected mortality rather than admission risk.

**Figure 1:**
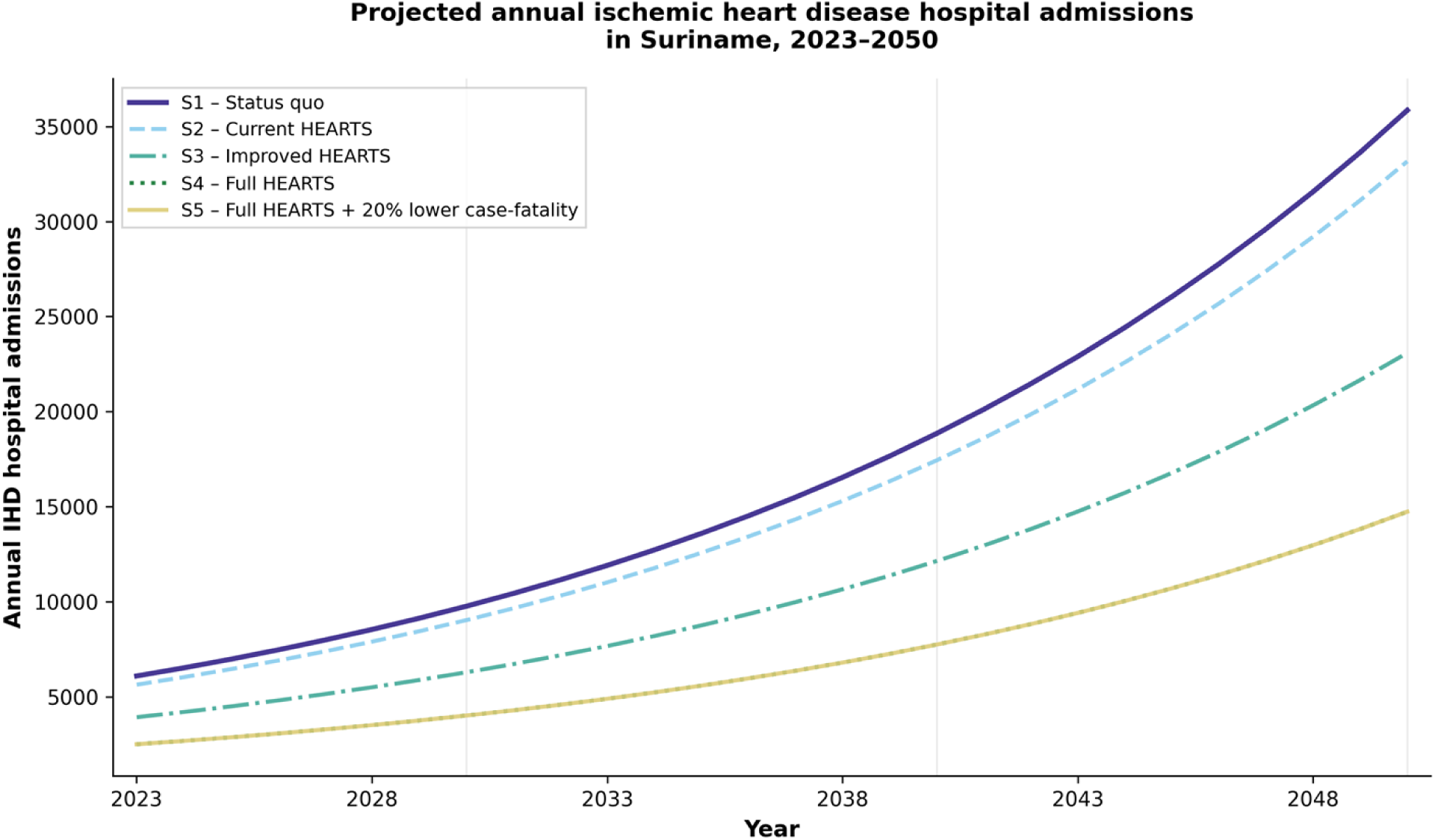
Projected annual ischemic heart disease hospital admissions in Suriname, 2023–2050. Deterministic projections of annual hospital admissions for ischemic heart disease (ICD-10: I20–I25) in Suriname from 2023 through 2050, stratified by five scenarios of cardiovascular prevention and health-system response. The status quo scenario (S1) projects admissions under observed historical trends (2013–2022) without intervention, resulting in more than a six-fold increase over the projection period. Scenarios S2 through S5 model the impact of the HEARTS (Hypertension, Elevated-risk, Awareness, Revised prescribing, Treatment targets, Structured follow-up) program at varying levels of implementation: S2 reflects current implementation (80% program reach, 52.6% treatment retention, 56.2% blood-pressure control), S3 represents improved HEARTS reach with enhanced follow-up retention (80%), S4 assumes full HEARTS scale-up with target-level blood-pressure control, and S5 applies the same admission reductions as S4 but with an additional 20% reduction in age- and disease-specific case-fatality rates to illustrate complementary gains from improved in-hospital acute care. Admissions are projected sequentially year-by-year from the 2022 baseline using age-specific annual percentage change (APC) coefficients derived from log-linear regression (2013–2022), applied with UN World Population Prospects 2024 annual population growth rates. Vertical reference lines indicate key planning milestones (2030, 2040, 2050). S4 and S5 trajectories overlap exactly for admissions because scenario S5 affects in-hospital mortality only, not admission demand (see Figure 3). The figure demonstrates that without intervention, Suriname faces substantial growth in cardiovascular hospital demand; full HEARTS implementation could reduce 2050 admissions by approximately 56% relative to status quo.

**Figure 2:**
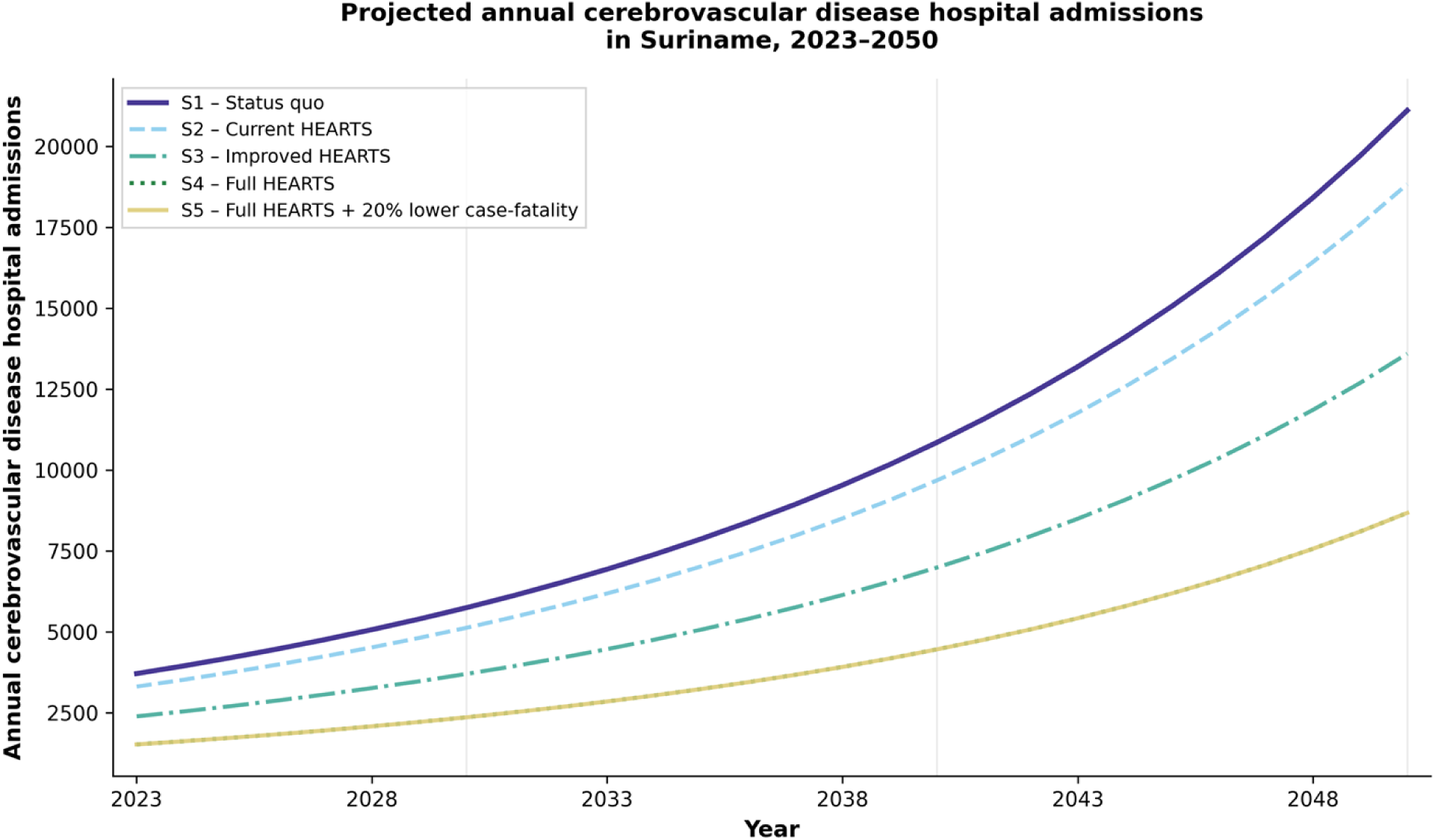
Projected annual cerebrovascular disease hospital admissions in Suriname, 2023–2050. Deterministic projections of annual hospital admissions for cerebrovascular disease (ICD-10: I60–I69; predominantly acute ischemic stroke and intracerebral haemorrhage) in Suriname from 2023 through 2050, under identical scenario definitions as Figure 1. Cerebrovascular disease shows steeper projected growth under the status quo scenario (S1) compared to ischemic heart disease, driven by higher age-specific annual percentage change (APC) coefficients in younger age groups, particularly ages 30–39 years (cerebrovascular APC: 12.73% vs ischemic heart disease APC: 9.25%). This age-pattern sensitivity is captured in the one-way sensitivity analysis presented in supplementary materials. As with Figure 1, scenarios S2–S4 progressively reduce admission volume through improved cardiovascular risk factor management and treatment adherence, while S5 applies the same admission trajectory as S4 with an additional 20% reduction in case-fatality rates to model improved hospital-based acute stroke care. Vertical reference lines indicate planning milestones (2030, 2040, 2050). S4 and S5 admissions overlap completely; mortality benefits of S5 are shown separately in Figure 3. Full HEARTS implementation (S4) could reduce 2050 cerebrovascular admissions by approximately 57% relative to status quo. The comparison between Figures 1 and 2 illustrates how disease-specific APC heterogeneity affects long-term projections and underscores the importance of age-stratified modelling for health-system planning.

**Table 2.** Projected annual cardiovascular hospital admissions under alternative policy pathways Policy pathway.

| Policy pathway | 2030 | 2040 | 2050 |
|---|---|---|---|
| S1. Status quo | 15,592 | 30,772 | 62,643 |
| S2. Current HEARTS | 14,5 | 28,516 | 58,073 |
| S3. Strengthened HEARTS | 10,352 | 20,367 | 41,69 |
| S4. Full HEARTS targets | 6,836 | 13,408 | 27,469 |
| S5. Full HEARTS + improved hospital survival | 6,836 | 13,408 | 27,469 |

*Values represent median probabilistic projections. Scenario differences were calculated within paired Monte Carlo simulations.*

### Projected in-hospital deaths

Under the status quo pathway, the increase in hospital admissions translated into a substantial increase in projected annual in-hospital deaths because age- and disease-specific CFRs were held constant. Projected deaths under status quo were 633 in 2030; 1,263 in 2040; and 2,644 in 2050. Increasingly intensive prevention produced progressively lower mortality. By 2050, projected deaths were: 2,644 under status quo; 2,452 under current HEARTS; 1,755 under strengthened HEARTS; 1,155 under full HEARTS; and 924 under full HEARTS plus improved hospital survival. The corresponding median reduction relative to status quo was approximately 7.3%, 33.6%, 56.3% and 65.0%, respectively. The difference between S4 and S5 demonstrates the distinct pathways through which prevention and hospital care operate. Both produced the same projected number of admissions, but S5 generated fewer deaths because of the additional illustrative CFR reduction.

**Table 3.** Projected annual in-hospital deaths under alternative policy pathways.

| Policy pathway | 2030 | 2040 | 2050 |
|---|---|---|---|
| S1. Status quo | 633 | 1,263 | 2,644 |
| S2. Current HEARTS | 590 | 1,173 | 2,452 |
| S3. Strengthened HEARTS | 432 | 837 | 1,755 |
| S4. Full HEARTS targets | 295 | 550 | 1,155 |
| S5. Full HEARTS + improved hospital survival | 236 | 440 | 924 |

### Policy differences at the 2050 planning horizon

By 2050, the divergence between the status quo and prevention pathways was substantial. Relative to status quo, median projected admissions were approximately 6.0% lower under current HEARTS; 33.5% lower under strengthened HEARTS; and 56.1% lower under full HEARTS. The corresponding uncertainty ranges were approximately 4.8–9.2%, 31.8–35.1%, and 54.7–57.7%, respectively. The full HEARTS plus improved hospital survival pathway produced the same admission reduction as full HEARTS but a larger reduction in mortality, with deaths approximately 65.0% lower than status quo in 2050. These results demonstrate that the model can distinguish between the potential effects of upstream prevention on hospital demand and downstream improvements in survival among patients who require hospital care.

**Figure 3:**
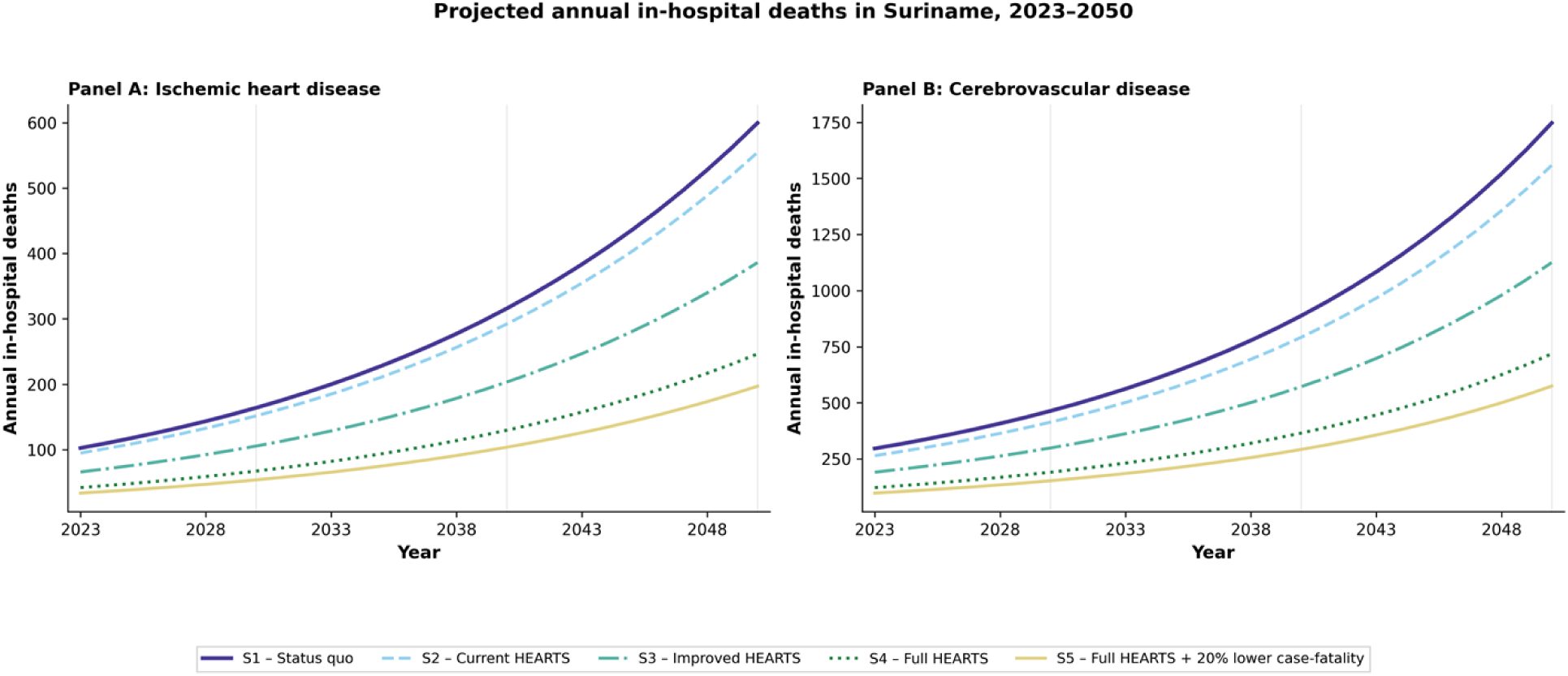
Projected annual in-hospital deaths in Suriname, 2023–2050. Two-panel deterministic projections of annual in-hospital deaths from cardiovascular disease in Suriname from 2023 through 2050. Panel A shows deaths from ischemic heart disease (ICD-10: I20–I25); Panel B shows deaths from cerebrovascular disease (ICD-10: I60–I69). All scenarios (S1–S5) are based on applying age- and disease-specific case-fatality rates (CFR) derived from 2022 hospital discharge data. For scenarios S1–S4, case-fatality rates are held constant throughout the projection period, allowing isolation of mortality burden attributable to changes in admission volume alone. Scenario S5 applies the same admission trajectory as S4 (identical to Figures 1 and 2) but incorporates an additional 20% relative reduction in age- and disease-specific CFR, representing an illustrative estimate of potential mortality gains from improved in-hospital acute care, post-acute management, and secondary prevention (e.g., thrombolysis, thrombectomy, intensive care protocols, discharge planning). This scenario is purely illustrative and not empirically grounded in observed data; it is included to demonstrate the complementary role of hospital-level survival improvements alongside population-level admission prevention. Under the status quo (S1), in-hospital deaths from both conditions more than double by 2050. S4 (full HEARTS) reduces 2050 deaths by approximately 56% for both diseases through admission prevention alone. S5 provides an additional 20% mortality reduction via CFR improvement, illustrating a potential pathway to more than 60% combined mortality reduction by 2050 if both prevention and acute-care quality are simultaneously advanced. Vertical reference lines denote planning milestones (2030, 2040, 2050). The divergence between S4 and S5 in Panel A and B is most pronounced at later projection years, reflecting the cumulative effect of the CFR reduction applied to large absolute numbers of admissions.

### Probabilistic uncertainty

The PSA demonstrated increasing uncertainty with increasing projection horizon. For the status quo pathway, the 2050 estimate of 62,643 admissions had a 95% UI of 46,351–102,113. The widening interval reflects uncertainty in the estimated historical age-specific trends and the compounding effect of these trends over time. The PSA used paired parameter draws across pathways, allowing scenario differences to be evaluated under the same underlying epidemiological uncertainty. The uncertainty analysis therefore provides information about the range of outcomes compatible with the specified model parameters rather than a probability distribution over all possible future cardiovascular trajectories.

### One-way sensitivity analysis

The OWSA identified several age- and disease-specific trends that strongly influenced projected outcomes. For admissions, the largest relative outcome range was associated with cerebrovascular disease among people aged 30–39 years (74.96%), followed by cerebrovascular disease among those aged 60–69 years (40.09%), IHD among those aged ≥70 years (27.85%), cerebrovascular disease among those aged ≥70 years (20.65%), and IHD among those aged 50–59 years (17.80%). For deaths, the largest relative ranges were observed for cerebrovascular disease among people aged 30–39 years (149.14%) and 60–69 years (64.19%), followed by cerebrovascular disease among people aged ≥70 years (52.53%), cerebrovascular disease among those aged 50–59 years (18.26%), and IHD among those aged ≥70 years (13.95%). The findings indicate that improvements in age-specific cardiovascular surveillance, particularly for cerebrovascular disease, could materially strengthen future decision-support models.

### Temporal holdout assessment

In the internal temporal holdout assessment, models fitted to 2013–2019 and used to estimate 2020–2022 demonstrated strong correspondence between observed and model-estimated values. Across the 36 age-disease-year combinations, the correlation was r=0.964, with R²=0.889, MAE of 117.5, RMSE of 179.7, and MAPE of 20.18%. The MAPE was 20.43% for IHD and 19.92% for cerebrovascular disease. These results support the usefulness of the trend framework for scenario analysis while also demonstrating meaningful prediction error. The temporal assessment therefore does not establish precise long-term predictive accuracy.

### COVID-19 exclusion analysis

Excluding the COVID-19 period from trend estimation had a limited effect on the long-term trajectory. The deterministic 2050 projection increased from approximately 56,959 admissions under the primary specification to approximately 57,643 admissions when the 2020–2022 period was excluded, corresponding to a difference of approximately 1.2%. Differences at earlier planning horizons were smaller. The findings suggest that the long-term status quo trajectory was not primarily driven by the inclusion of the pandemic years.

### Discussion Principal findings

This study demonstrates how routinely collected hospital data can be transformed into a quantitative framework for cardiovascular health-system planning. Three findings are particularly important. First, if historical admission trends continue, Suriname could experience a substantial increase in annual cardiovascular hospital demand. The status quo pathway projected 62,643 annual IHD and cerebrovascular disease admissions by 2050, with a 95% UI of 46,351–102,113. Second, alternative prevention pathways produced substantially different trajectories. By 2050, projected admissions were approximately 6% lower under current HEARTS, 34% lower under strengthened HEARTS and 56% lower under the full HEARTS pathway. Third, prevention and hospital care address different components of the health-system problem. Prevention primarily changes the number of cardiovascular events reaching hospital, whereas improved hospital survival changes the probability of death among patients who are hospitalized. The combined pathway therefore generated the lowest projected mortality(14). The principal contribution of the study is consequently not a prediction of the exact number of cardiovascular admissions in 2050. Rather, it is the demonstration of how epidemiological surveillance can be translated into policy-relevant alternative futures.

### From routine health data to actionable decision support

A central contribution of this study is to demonstrate how routinely collected hospital data can be transformed into actionable evidence for public-health policy and health-system planning(4). In settings such as Suriname, where comprehensive population-based disease registries may be limited, routinely generated hospital information can nevertheless provide an important basis for identifying emerging patterns in cardiovascular disease and anticipating future service needs. By combining observed age- and disease-specific admission trends with population projections and alternative prevention and cardiovascular-care scenarios, this study moves beyond describing historical burden to examining how different policy pathways could shape future hospital demand(15). This approach is particularly relevant to health-system strengthening in the Americas, where policymakers must allocate limited resources across prevention, primary care, chronic disease management, and hospital services(16). The projections should not be interpreted as precise forecasts of future admissions or as estimates of the causal effect of HEARTS. Rather, they quantify alternative trajectories conditional on explicit assumptions and therefore provide a transparent framework for policy discussion and planning. The broader implication is that routine health information systems do not need to be perfect or fully population-based to contribute to strategic planning; when their coverage and limitations are clearly defined, they can be linked to modelling approaches that generate decision-relevant evidence for national and local health authorities(17).

### Prevention as a health-system strategy

The results also highlight prevention as an integral component of health-system planning rather than a policy domain separate from hospital capacity(18). Under the status quo scenario, admissions for ischemic heart disease and cerebrovascular disease increased substantially toward 2050, whereas progressively stronger prevention and cardiovascular-care pathways were associated with markedly lower projected demand. By 2050, the modelled scenarios corresponded to approximately 6%, 34%, and 56% fewer admissions than the status quo under current, strengthened, and full HEARTS implementation, respectively. These estimates are counterfactual scenario results rather than observed program effects. Nevertheless, they demonstrate the potential scale of the relationship between population-level cardiovascular prevention and downstream health-service demand(19). For health systems with constrained resources, preventing avoidable cardiovascular events can therefore be viewed simultaneously as a public-health objective and a capacity-management strategy. The findings support a health-system approach in which investments in primary prevention, hypertension detection and control, treatment continuity, and cardiovascular risk management are considered alongside investments in emergency and inpatient care. Such integration is consistent with the broader objective of strengthening health systems by reducing preventable demand while ensuring that adequate capacity remains available for patients who develop acute cardiovascular disease(20).

### HEARTS implementation as a modifiable health-system pathway

The scenario analysis provides a practical illustration of how implementation gaps within a population hypertension program can be translated into potential long-term health-system consequences. The Surinamese HEARTS program has established a national implementation platform involving primary-care facilities and health workers, but available program data indicate that important gaps remain in follow-up and blood-pressure control (10). In the present model, strengthening treatment and follow-up retention and achieving the predefined implementation targets resulted in progressively lower projected cardiovascular hospital demand. These findings should not be interpreted as an evaluation of HEARTS effectiveness, because the model combines observed program indicators with assumptions about future program reach, retention, and control. In particular, the assumed 80% reach represents a policy scenario rather than observed national coverage. The value of the analysis is instead that it makes implementation ambition measurable in terms of potential downstream demand. This creates a direct link between program monitoring and health-system planning: indicators such as program reach, retention, treatment continuity, and blood-pressure control can be followed not only as measures of primary-care performance but also as leading indicators of future cardiovascular burden(21). For PAHO Member States pursuing integrated hypertension and cardiovascular disease strategies, this type of framework may help connect implementation monitoring with longer-term resource planning(22).

### Hypertension control as the prevention–hospital interface

Hypertension provides a particularly important link between primary-care performance and hospital demand because inadequate blood-pressure control contributes substantially to both ischemic heart disease and cerebrovascular disease. The scenario framework incorporated published evidence from 36 countries in the Americas showing an association between population hypertension control and mortality from ischemic heart disease and stroke(12). However, this ecological evidence does not directly estimate the effect of HEARTS implementation on hospital admissions in Suriname and cannot establish a causal pathway from program implementation to hospital utilization. It was therefore used as an external parameter within counterfactual scenarios rather than as a direct estimate of program impact. This distinction is important, but it does not diminish the policy relevance of the analysis. Instead, it identifies a measurable pathway through which improvements in hypertension management could potentially influence downstream cardiovascular burden(23). For Suriname, and potentially other health systems in the Americas facing similar data constraints, strengthening routine measurement of hypertension treatment, retention, control, cardiovascular events, and hospital utilization would allow future analyses to replace externally derived associations with locally estimated relationships. In this way, the model can also serve as a starting point for strengthening the information systems required to evaluate and improve cardiovascular programs.

### Younger adults and the need for a life-course prevention approach

The projected trends among younger adults provide an additional public-health signal that is particularly relevant for prevention policy. The highest estimated annual percentage increase in ischemic heart disease occurred among adults aged 30–39 years, while cerebrovascular disease also showed its largest estimated increase in this age group(24,25). Although older adults continue to account for a large proportion of absolute cardiovascular admissions, rapidly increasing rates among younger adults suggest that cardiovascular prevention should not focus exclusively on populations already approaching older age(26). Earlier cardiovascular disease may also have wider social consequences because affected individuals are more likely to be economically active and may experience prolonged disability or loss of productivity. These findings therefore support a life-course approach to cardiovascular risk reduction, including earlier identification and management of hypertension and other modifiable risk factors(27). At the same time, age-specific uncertainty needs to be considered: some estimated trends were not statistically distinguishable from no change. The policy implication is consequently not that every age-specific projection should be treated as certain, but that prevention strategies should include younger and middle-aged adults rather than relying primarily on interventions delivered after cardiovascular risk has already accumulated. This is particularly relevant to health-system strengthening because prevention initiated earlier in the life course may reduce future demand while simultaneously improving population health(25,27,28).

### Implications for health-system capacity and service planning

The projected increase in cardiovascular admissions has implications across the continuum of care and should not be translated simply into a requirement for additional hospital beds. Increasing cardiovascular demand may place pressure on emergency departments, diagnostic imaging, cardiology and neurology services, interventional procedures, inpatient nursing, rehabilitation, medicines, and post-discharge follow-up(29). The divergence between the modelled scenarios indicates that future service requirements will depend partly on the effectiveness and scale of prevention and chronic disease management. This supports a coordinated health-system planning approach in which primary-care strengthening and hospital preparedness are developed together(29). Importantly, the findings do not imply that prevention can substitute for investment in acute cardiovascular services. Even under the most effective modelled prevention pathway, substantial cardiovascular demand remains. Rather, prevention can reduce avoidable demand while health systems maintain sufficient capacity for patients who continue to experience acute events(30). The results may therefore assist health authorities in considering where additional investments are likely to have the greatest long-term system value, including hypertension control, continuity of chronic care, emergency cardiovascular pathways, diagnostic capacity, specialist workforce, and rehabilitation. Because the underlying data cover three major public hospitals and approximately 71.8% of overall hospital admissions during the study period, the projected absolute numbers should not be interpreted as a complete national estimate of service requirements. Their principal value for policy is to demonstrate the potential direction and relative scale of future demand and the extent to which alternative prevention pathways could modify it.

### The importance of acting before demand peaks

The widening differences between scenarios over time have an important implication for public-health policy: decisions made during the current decade may influence the health-system pressures experienced decades later. The divergence between the status quo and higher-implementation scenarios was already evident by 2030 and became substantially larger by 2040 and 2050. This temporal pattern is relevant because strengthening primary care, improving hypertension control, developing the cardiovascular workforce, and expanding hospital capacity all require time to implement. Delaying action until demand reaches its projected maximum may therefore constrain the ability of the health system to respond efficiently. At the same time, the projections should not be interpreted as certainty about the level of demand in 2050. The temporal holdout analysis demonstrated good agreement between observed and predicted admission patterns but also meaningful prediction error, and the probabilistic analysis showed increasing uncertainty over longer projection horizons. The most appropriate policy use of these findings is therefore not to plan a fixed system around a single 2050 estimate, but to use 2030, 2040, and 2050 as planning horizons and reassess trajectories as new data become available. This supports a proactive rather than reactive approach to cardiovascular health-system strengthening.

### Adaptive planning and learning health systems

The uncertainty surrounding long-term cardiovascular demand further supports an adaptive approach to health-system planning. In the present analysis, uncertainty increased substantially toward 2050, and the one-way sensitivity analysis identified specific age- and disease-specific trends that contributed disproportionately to variation in projected outcomes. For example, the trend for cerebrovascular disease among adults aged 30–39 years produced the largest relative range for projected admissions, while cerebrovascular trends in several other age groups also contributed substantially to uncertainty. These findings have practical implications for surveillance priorities: improving the quality and completeness of age-specific cardiovascular data could directly strengthen future planning. Rather than treating the projections as a one-time forecasting exercise, health authorities could use the framework as part of an iterative planning cycle in which hospital trends, hypertension-control indicators, HEARTS implementation, and health-system capacity are periodically reassessed and projections updated. Such an approach is consistent with a learning health-system model in which routine data are continuously converted into information for policy adaptation(31,32). The value of the framework therefore lies not in eliminating uncertainty, but in making uncertainty explicit, identifying the parameters that matter most, and creating a mechanism through which improved surveillance and program implementation can progressively improve future decision-making.

### Strengths

This study has several strengths that enhance its relevance for public-health policy and health-system planning. First, it uses a decade of routinely collected hospital data and disaggregates cardiovascular admission trends by disease and age group, allowing heterogeneous trajectories to be identified rather than assuming a uniform cardiovascular trend. Second, the framework integrates observed hospital trends with projected population growth and explicitly defined prevention and cardiovascular-care pathways. Third, the analysis links epidemiological projections to a real-world health-system intervention, HEARTS, while clearly distinguishing observed program indicators from modelling assumptions. Fourth, uncertainty was examined using both probabilistic sensitivity analysis and one-way sensitivity analysis, allowing the overall uncertainty of the projections and the parameters most influential to outcomes to be identified. Fifth, temporal holdout assessment provided an internal assessment of model performance against more recent observed admission patterns, while exclusion of the COVID-19 period assessed the influence of pandemic-related disruption on the long-term trend estimates. Finally, the framework is transparent enough to be updated as new routine health data and program indicators become available. This combination of routine surveillance, scenario modelling, and implementation-oriented interpretation provides a potentially transferable approach for other health systems in the Americas where data are incomplete but policy decisions nevertheless require forward-looking evidence.

### Limitations

Several limitations should be considered when interpreting these findings. First, the analysis was based on hospital discharge data from three major public hospitals and therefore does not constitute a complete national population-based registry. Although these hospitals represented approximately 71.8% of overall hospital admissions during the study period, disease-specific coverage may differ, particularly because cardiovascular admissions also occur in private hospitals. Second, the unit of analysis was the hospital admission rather than the unique patient; individuals with multiple admissions could therefore contribute more than once, and transfers between participating hospitals may result in more than one admission being recorded for a single clinical episode. Third, disease classification was based on discharge diagnoses and may be influenced by diagnostic and coding practices. Fourth, the historical trend model assumes that past age- and disease-specific trends provide an informative basis for long-term extrapolation, although future epidemiological, demographic, treatment, and health-system conditions may differ from those observed during 2013–2022. Fifth, the population projections were derived from UN-projected annual population growth rates and applied sequentially across age groups because age-specific annual growth rates were not used. Sixth, the status quo mortality scenario assumes that 2022 age- and disease-specific case-fatality rates remain constant throughout the projection period.

Seventh, the HEARTS scenarios combine observed program indicators with assumptions about future program reach, treatment/follow-up retention, and blood-pressure control; incomplete program reporting means that these indicators should not be interpreted as nationally representative estimates. Eighth, the assumed 80% program reach and the 20% case-fatality reduction in Scenario 5 are illustrative modelling assumptions rather than empirically established national effects. Ninth, the external hypertension-control associations were derived from an ecological study across countries in the Americas and were not specific to Suriname, HEARTS implementation, or hospital admissions. Finally, the PSA captures uncertainty in the stochastic parameters included in the model but does not encompass all forms of structural uncertainty, including future changes in disease incidence, treatment patterns, hospital utilization, program implementation, coding practices, or unforeseen health-system shocks. The projections should therefore be interpreted as scenario-based decision-support estimates rather than precise predictions of national cardiovascular admissions or deaths.

### Conclusion

Suriname may face a substantial increase in cardiovascular hospital demand if the admission trends observed between 2013 and 2022 continue. Under the status quo pathway, combined annual IHD and cerebrovascular disease admissions were projected to reach 62,643 by 2050, with a 95% uncertainty interval of 46,351–102,113. The modelled policy pathways demonstrated that progressively stronger cardiovascular prevention could substantially alter this trajectory. Under the full HEARTS pathway, projected 2050 admissions were approximately 56% lower than under status quo. An additional illustrative improvement in hospital survival further reduced projected deaths without changing the admission trajectory. These findings should not be interpreted as precise forecasts or causal estimates of HEARTS effectiveness. Their principal value lies in demonstrating how routinely collected health data can be transformed into a transparent, policy-relevant decision-support framework. For Suriname, the findings support an integrated approach in which sustained investment in hypertension control and cardiovascular prevention is combined with continued strengthening of acute cardiovascular and cerebrovascular care. More broadly, the study illustrates how health systems with limited resources can use routine epidemiological data not only to describe current disease burden, but also to explore alternative futures and inform decisions about prevention, capacity, workforce, medicines, surveillance and resource allocation. Suriname’s future cardiovascular hospital burden is therefore not predetermined. The trajectory will depend in part on the policy choices, implementation capacity and health-system investments made today.

## Declarations Ethics approval

Ethical approval for the hospital-data analyses was obtained from the Commissie Mensgebonden Onderzoek (CMWO), Suriname, letter CMWO-14, dated 25 March 2026. The study used anonymized routinely collected health data, and informed consent was waived in accordance with the applicable national framework.

## Data availability statement

The underlying hospital data are not publicly available because of restrictions imposed by the Ministry of Health of Suriname. Data may be made available upon reasonable request and with appropriate permission from the Ministry of Health. This wording is consistent with the data-availability statement used elsewhere in the research program.

## Author contributions statement

**RAD** conceived the study, interpreted the data, created the figures, and drafted the manuscript. **JRT and RB** provided overall scientific guidance and supervision. **RB** contributed to manuscript editing. **RGS** provided institutional support, feedback, and permission to conduct the study. **AM and VJ** supported data acquisition and provided critical feedback. **MB** contributed to the critical review of the manuscript, including validation of numerical findings and input on the strengths and limitations of the study. All authors critically reviewed the manuscript, approved the final version, and agree to be accountable for all aspects of the work.

## Declaration of Use of Artificial Intelligence

Artificial intelligence (AI) tools were used during the preparation of this manuscript. Claude (Anthropic) was used to support the model sensitivity analysis and the preparation of study figures. ChatGPT (OpenAI) was used to support text editing, grammar correction, and language refinement. The authors designed the study, determined the analytical approach and modelling assumptions, verified the analyses and results, and reviewed the figures and manuscript. All AI-assisted outputs were critically reviewed and edited by the authors as appropriate. The authors remain fully responsible for the accuracy, integrity, interpretation, and presentation of the work, including all analyses, figures, and conclusions.AI tools were used as supportive tools and did not replace the authors’ scientific judgment or responsibility for the research.

